# The COVID-19 herd immunity threshold is not low: A re-analysis of European data from spring of 2020

**DOI:** 10.1101/2020.12.01.20242289

**Authors:** Spencer J. Fox, Pratyush Potu, Michael Lachmann, Ravi Srinivasan, Lauren Ancel Meyers

## Abstract

The recent publication of the Great Barrington Declaration (GBD), which calls for relaxing all public health interventions on young, healthy individuals, has brought the question of herd immunity to the forefront of COVID-19 policy discussions, and is partially based on unpublished research that suggests low herd immunity thresholds (HITs) of 10-20%. We re-evaluate these findings and correct a flawed assumption leading to COVID-19 HIT estimates of 60-80%. If policymakers were to adopt a herd immunity strategy, in which the virus is allowed to spread relatively unimpeded, we project that cumulative COVID-19 deaths would be five times higher than the initial estimates suggest. Our re-estimates of the COVID-19 HIT corroborate strong signals in the data and compelling arguments that most of the globe remains far from herd immunity, and suggest that abandoning community mitigation efforts would jeopardize the welfare of communities and integrity of healthcare systems.

## Commentary

The time course and burden of the COVID-19 pandemic will depend on the herd immunity threshold (HIT) of the virus, which is the fraction of the population that needs to be immunized for an epidemic to slow in the absence of mitigation efforts. Estimates for the COVID-19 HIT range from 6% to over 60%^1,2^. Given that roughly 10% of the global population has been infected^3^, the low end of this range implies that the pandemic should soon burn out on its own, while the high end paints a grim picture of future morbidity and mortality, in the absence of pervasive non-pharmacological interventions, efficacious vaccines, or life-saving drugs.

The recent publication of the Great Barrington Declaration (GBD), which calls for relaxing all public health interventions on young, healthy individuals, has brought the question of herd immunity to the forefront of COVID-19 policy discussions^4,5^. The authors state that “immunity in the population is playing a substantial role in controlling the spread,” tacitly referencing preprints by multiple GBD authors that posit HITs of 10-20%^2,6^. Evidence against this claim is mounting, including pandemic resurgences throughout Europe and the US and attack rates exceeding 50% in the hardest hit regions and congregate living settings^7–9^.

Given that this unpublished work is fundamentally shaping public discourse and global policy, reconciling its claims with the rapidly evolving state of the pandemic is paramount. To this end, we reevaluated the core model from the study and have identified a fundamental flaw that leads to underestimation of the COVID-19 HIT. The authors sought to identify the *cause* of the summer slowdown in four European countries by *fitting* an SEIR-like model of COVID-19 transmission to case count data up to July of 2020. The analysis is structured so that one of two explanations are possible. Either the pandemic is self-limiting (i.e., the HIT is low) or social distancing and other community mitigation efforts slowed transmission. However, teasing apart the contributions of these factors from the case data alone is statistically impossible. In other words, one cannot estimate the HIT without making assumptions about the efficacy of community mitigation, and vice versa (See supplement).

So the researchers made a strong assumption about community mitigation efforts in Europe in the spring and summer of 2020. Roughly, they assume that Europe locked down throughout April and then returned to *normal* (linearly) by the end of August (Figure 1A - Blue). By assuming that interventions disappear steeply, the model concludes that the pandemic must be fading due to immunity buildup, and thus estimates low HITs. As it turns out, the derived HIT is highly sensitive to the assumed timeline of mitigation (Figure S2-S3) and we have good reason to believe their assumption is flawed. The authors use mobility traces to justify their pattern^2^, but other precautionary policies like school closures, wearing of face coverings, and social distancing have likely kept transmission repressed far below the pre-April baseline (Figure 1A - Black). When we plug this *plausible* scenario into the Aguas et al. model^2^ (Figure 1A - Green), the COVID-19 HIT estimate increases six-fold for Belgium, three-fold for England, ten-fold for Portugal, and six-fold for Spain (Figure 1B). A range of alternative scenarios produce similar estimates (Figure S2-S3). If policymakers were to adopt a *herd immunity strategy*, in which the virus is allowed to spread relatively unimpeded, we project that cumulative COVID-19 deaths would total almost 650,000 (95% CI: 500,000 - 780,000) across all four countries through the end of the pandemic under the revised HIT estimates, roughly five-fold higher that projected under the original low HIT estimates (Figure S4).

**Figure 1:**
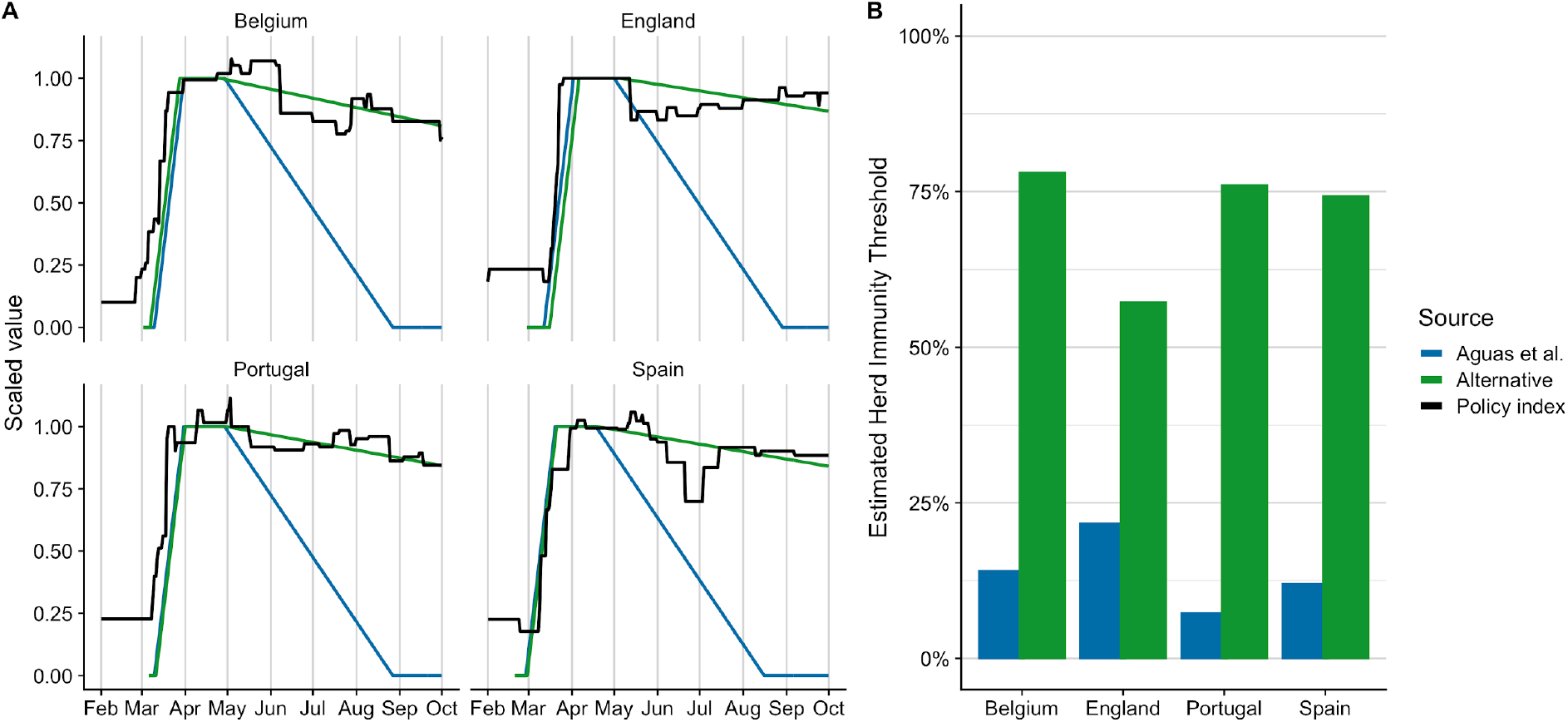
Re-estimation of the COVID-19 herd immunity thresholds (HIT) in four European countries, using the model of Aguas et al.^2^. (A) Strength of non-pharmacological interventions indicated by a government response index^11^ (black) compared to the trends assumed by Aguas et al.^2^ (blue) and *plausible* alternatives (green) derived to match the observed indices. The time-series are scaled for comparison, where values of zero and one correspond to lack of mitigation efforts and the average level of community mitigation in April, respectively. (B) Estimated HIT using the Aguas et al.^2^ approach, under the original assumption that non-pharmacological efforts rapidly decrease to baseline (blue) and alternative assumption that measures mirror the government response index (green).

The fragility of the Aguas et al. study^2^ undermines a key premise of GBD and other recent calls for “herd immunity” strategies. To their credit, the authors clearly demonstrate that population heterogeneity in susceptibility to infection can dramatically lower the herd immunity threshold^10^. However, their model can only disentangle the impacts of heterogeneity versus interventions on COVID-19 transmission when approached with sufficient data and validated assumptions. Our rough, but arguably more plausible, re-estimates of the COVID-19 HIT corroborate strong signals in the data and compelling arguments that most of the globe remains far from herd immunity. Moreover, abandoning community mitigation efforts would jeopardize the welfare of communities and integrity of healthcare systems.

## Data Availability

All code and data are available at https://github.com/pratyush16/VariationalSusceptibility

## Acknowledgements

The authors thank the authors of Aguas et al.^2^, for sharing their research code and data, which allowed us to replicate and extend their analyses. We acknowledge the UT COVID-19 Modeling Consortium for helpful discussion and comments, and the Texas Advanced Computing Center (TACC) at The University of Texas at Austin for providing HPC resources that have contributed to the research results reported within this paper. URL: http://www.tacc.utexas.edu. We also acknowledge support from a CDC COVID-19 supplementary grant (U01IP001136-01-01).

## Author Contributions

SJF, ML, and LAM conceived of the initial idea. PP carried out all analyses. RS and ML developed the analytical framework for statistical identifiability. SJF and LAM supervised the research. SJF and LAM wrote the initial draft. All authors significantly contributed to the manuscript and approved the final draft. PP, SJF, and RS verified the underlying data.

## Supplementary information

### 1 Overview

The low COVID-19 herd immunity thresholds (HITs) of 6-21% estimated in Aguas et al. [1] are inconsistent with other model-derived estimates [5, 9, 2] and seroprevalence-based estimates from some of the hardest hit regions around the world [3]. To explain this apparent discrepancy, we conducted a thorough review of their methods and code. We identified key assumptions about the timing and extent of community mitigation efforts that shift the COVID-19 HIT estimates downward. As their code was made openly available (https://github.com/mgmgomes1/covid), we apply their exact model structure and fitting procedure to evaluate the sensitivity of the HIT estimates to these assumptions.

Below, we detail our: (1) slight modification to their model fitting procedure, (2) sensitivity analyses with respect to the assumed mitigation curves (i.e., timing and magnitude of transmission reduction via non-pharmacological interventions), (3) derivation of alternate *plausible* mitigation curves from the Oxford COVID-19 Government Response Tracker response index data, (4) long-range COVID-19 mortality projections depending on the estimate HIT, and, finally (5) mathematical argument regarding the statistical non-identifiability the model (i.e., inability to simulataneously estimate the impact of community mitigation and population heterogeneity). The code used for the primary analyses along with the data can be accessed here: https://github.com/pratyush16/VariationalSusceptibility

### 2 Modification to the Aguas et al. model

We slightly modified the modeling framework of Aguas et al. [1] to include an additional month of COVID-19 incidence data (through August 7th, 2020) for Belgium (https://epistat.wiv-isp.be/covid/) and England (https://coronavirus.data.gov.uk/details/about-data). For Portugal and Spain we analyzed data through July 10th, 2020 as in [1]. We made this adjustment to address a strong tendency of the model to estimate a low HIT without evidence of increased transmission following intense mitigation during the initial stay at home orders.

### 3 Sensitivity of COVID-19 HIT estimates to mitigation curves

We analyzed the sensitivity of the HIT estimates from Aguas et al. to variations in the assumed temporal progression of the mitigation curves within their modeling framework. Specifically they assume a general shape of mitigation where mitigation begins, increases until it reaches a maximum level, remains at the maximum for some time, and then slowly returns to a baseline of no mitigation. In this shape there are five key parameters governing the progression: (1) the time that mitigation begins, (2) the time it takes for mitigation to take full effect, (3) the maximum impact of mitigation, (4) the duration that mitigation remains at maximum, and (5) the time for mitigation to be completely removed.

The modeling framework fits the timing that mitigation begins and the maximum impact it will have, but makes assumptions about the remaining parameters. Their framework assumes that once mitigation begins it will take 21 days to reach a maximum level, will remain there for 30 days, and will subsequently return to the original baseline level after 120 days. They test sensitivity to slight deviations of the time to return to baseline and find minimal change in HIT estimates looking at 150 or 180 days. As noted in the manuscript and seen in Figure 1, we believed their assumptions about mitigation progression were driving their low HIT estimates, so we tested the sensitivity of the low HIT estimates to a wide-range of values for the maximum mitigation duration (Figure S2) and times to return back to baseline (Figure S3). It’s clear that the model is sensitive to the assumed shape of mitigation, with HIT estimates ranging from almost 5% to 85% depending on the country and assumed shape.

It is clear that the estimated HITs are extremely sensitive to these assumptions, and that there are many combinations of heterogeneity and mitigation progression that can give similar fits (described in the next section), so it is extremely important that the assumed mitigation curves match reality. While Aguas et al. matched their mitigation shapes to mobility data, these data are not an accurate picture for total transmission mitigation, as decoupling of mobility and transmission has been previously noted as populations adopt precautionary behavior like mask wearing and social distancing [12].

Instead, we focused on the government response index developed by researchers at the University of Oxford [7]. This index captures 18 policy indicators widely implemented around the world, and produces an overall value that captures the strength of government responses through time as shown in Figure 1A. We found that the Aguas et al baseline assumptions did not match the actual progression of mitigation policies, and instead chose to fit the estimated time to completely remove mitigation according to the government response index. We assumed the baseline assumptions of 21 days to implement mitigation and 30 days at maximum mitigation impact, and fit the timing to completely remove mitigation based on the trends in the government response index. Fitted estimates for the time to return to normal can be found in Figure S3 for each country, and are almost an order of magnitude larger than assumed in Aguas et al (120): Belgium (828 days), England (1,130 days), Portugal (981 days), and Spain (1,033 days). While the government response index also likely doesn’t capture the full progression of community mitigation, because it doesn’t capture adherence to policies, we feel that it likely captures general countrywide trends better than mobility data alone.

**Figure S1:**
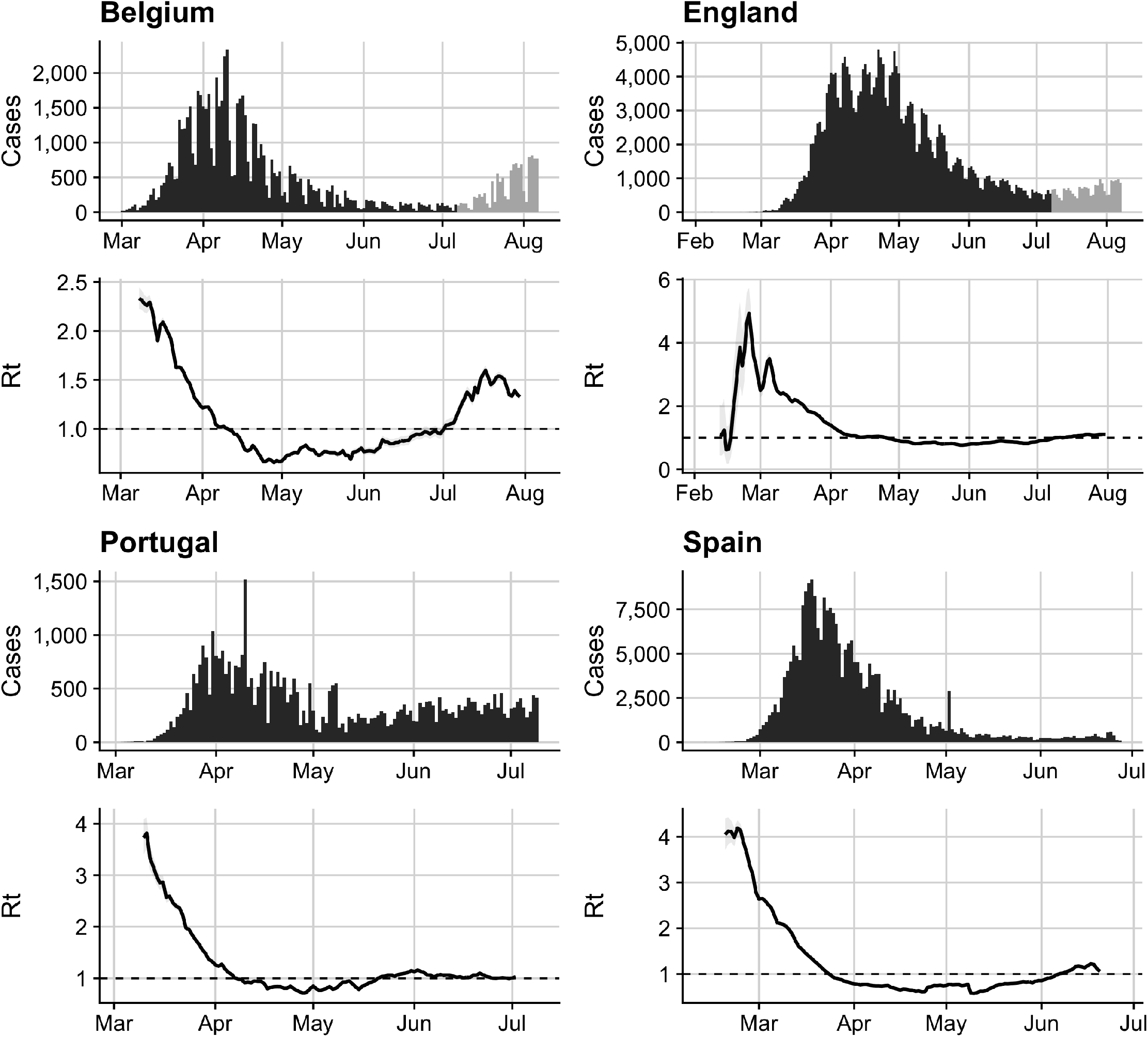
Country-specific case counts used for fitting alongside daily estimates of the effective reproduction number. Case counts in black indicate data used that are the same as Aguas et al, and light grey indicates the additional data we included in our fitting procedure as described in the modifications section of the supplement.

**Figure S2:**
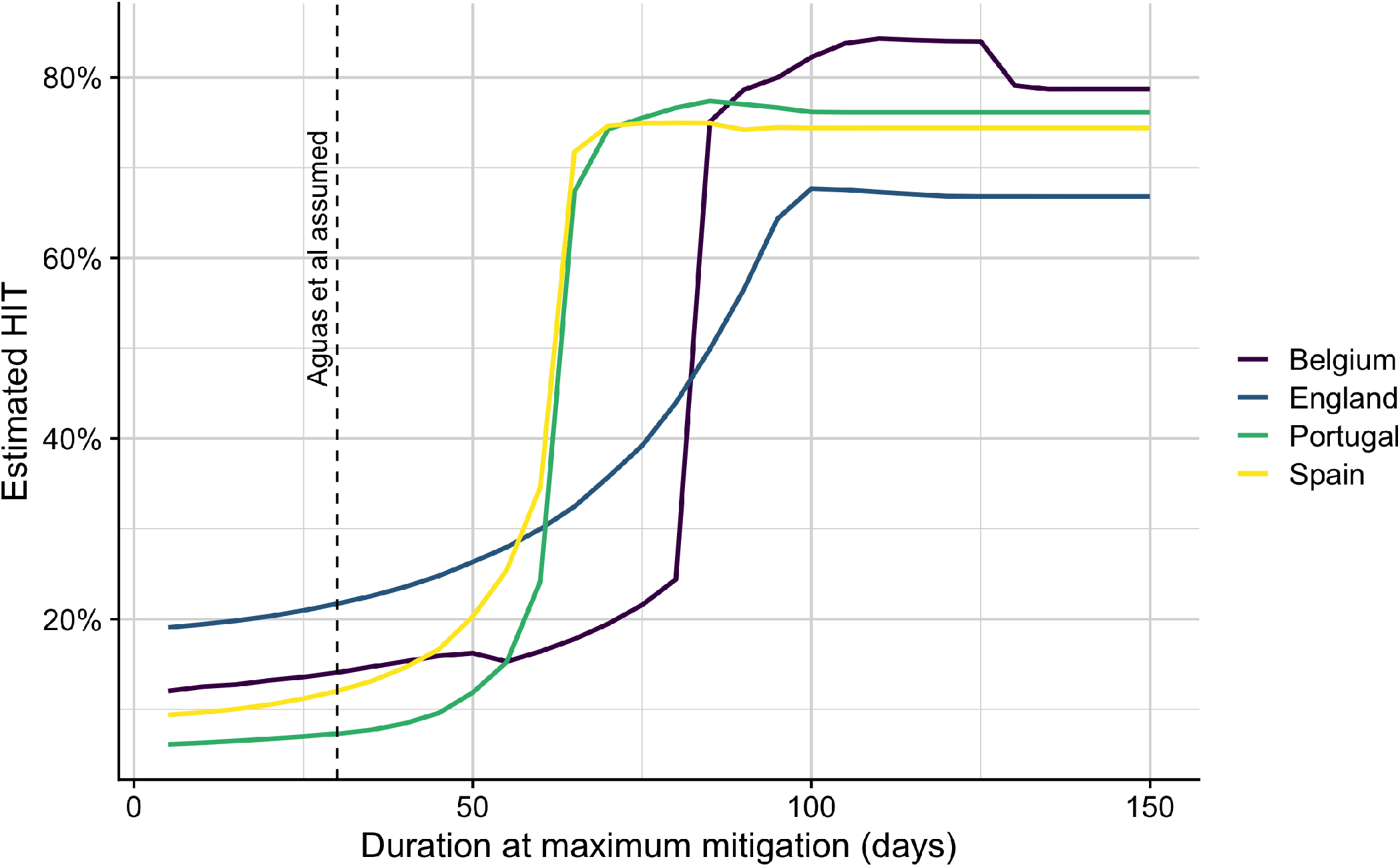
Sensitivity of herd immunity threshold (HIT) estimates to assumptions about the duration at maximum mitigation. Vertical black dashed line indicates the assumed value in Aguas et al. All estimates are made assuming the baseline value of time for mitigation to be completely removed of 120 days.

### 4 Projecting mortality under a *herd immunity* policy

We calculated the final epidemic size for an uncontrolled epidemic based on the method described in [6] for each country. We carried out the same fitting procedure as Aguas et al. using either their assumed mitigation curve or our revised version, we then calculated the mortality cost of following a “herd immunity strategy” using the estimated parameters from each European country. For each European country we used the estimated final epidemic size estimated for that scenario as the total infection count and an infection fatality rate of 0.68% (95% CI: 0.53%-0.82%) [10]. For the US, we assumed average final epidemic sizes based on the average of the estimates for the four countries for each assumed mitigation curve scenario, which suggests final epidemic sizes of 13.9% and 79.2% for the Aguas et al. and alternative scenarios respectively. Estimates for both scenarios can be seen in Figure S4. Country population sizes were assumed to be 56,286,961 (England), 11,607,113 (Belgium), 10,186,314 (Portugal), 46,761,086 (Spain), and 331,674,530 (United States) as estimated from UN population statistics made available through hrefhttps://www.worldometers.info/.

**Figure S3:**
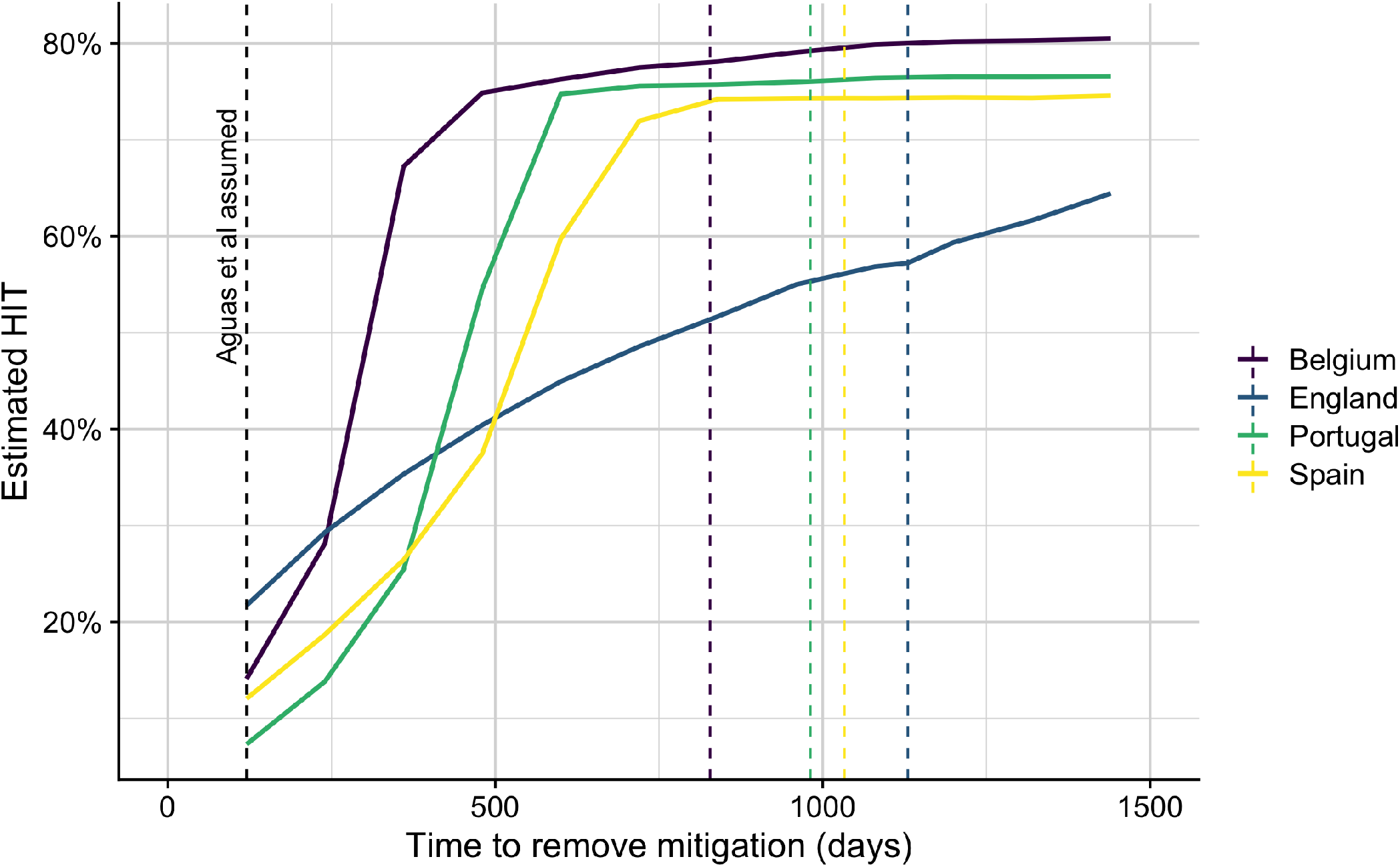
Sensitivity of herd immunity threshold (HIT) estimates to assumptions about the time for mitigation to be completely removed. Colored solid lines show sensitivity across all four European countries. Vertical black dashed line shows the assumed value in Aguas et al, and vertical colored lines show the fitted value from government response index used in our main analysis for each of the specific countries. All estimates are made assuming the baseline value of maximum mitigation duration of 30 days.

**Figure S4:**
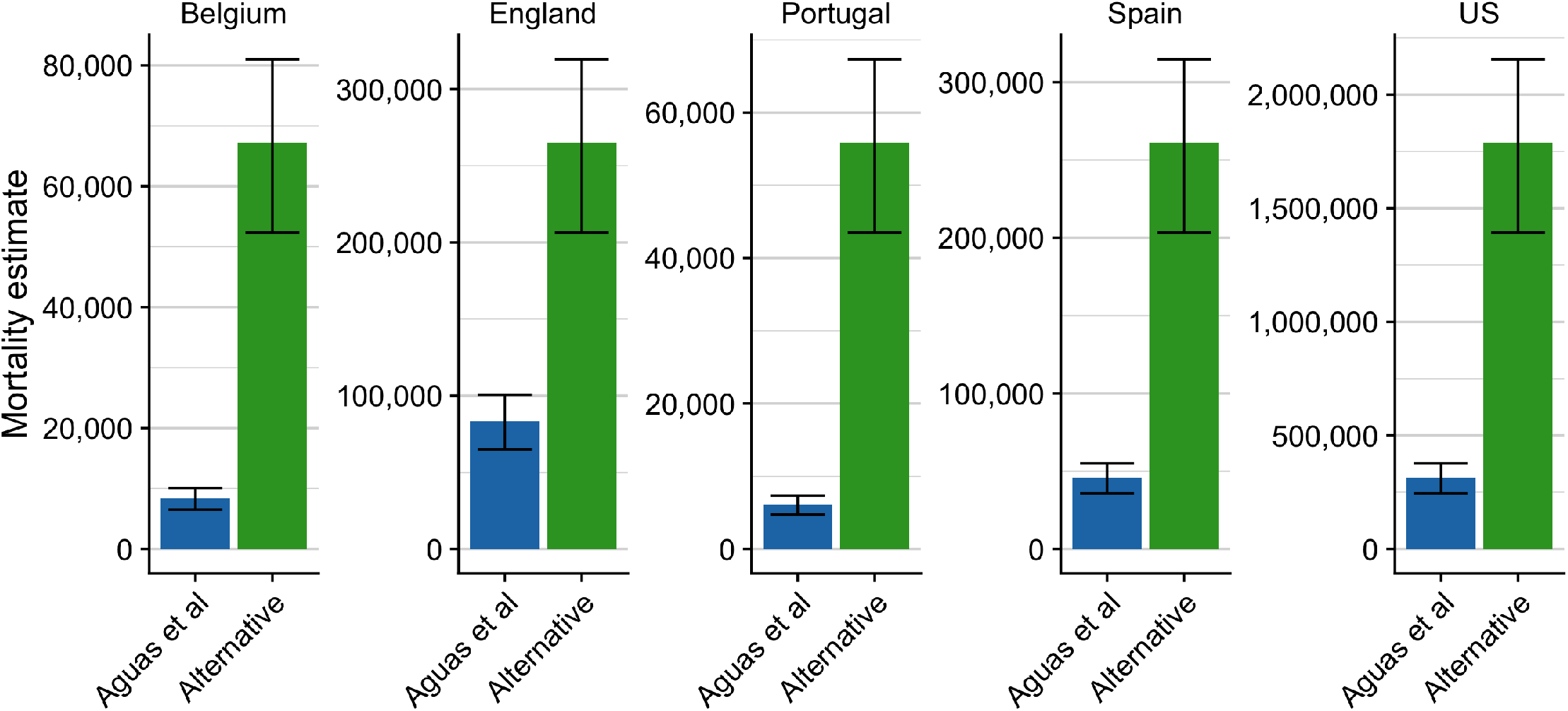
Mortality estimate for a herd immunity strategy for each of the four European countries and the United States. Comparison between estimated mortality if herd immunity thresholds are as low as estimated in Aguas et al (Blue), versus those estimated using the policy-based, alternative mitigation curve (Green). Estimates assume an infection fatality rate of 0.68% (95% CI: 0.53%-0.82%) [10]

### 5 Non-identifiability of the model

The model fitting procedure from Aguas et al. explains transmission dynamics between mitigation and population heterogeneity, which impacts herd immunity thresholds. In Aguas et al., they begin with assumed mitigation curves and estimate the herd immunity threshold. Here we describe mathematically why their estimation procedure is only identifiable with strong assumptions about the shape of the mitigation curves. We first break down the model structure into component parts to make clear the tension between mitigation and herd immunity. Then we use this analytical framework to flip their estimation procedure around to show how one can estimate mitigation curves that fit epidemic trajectories for any assumed herd immunity threshold.

To start, consider an SEIR model with variable susceptibility *S*_*t*_(*x*) and time-dependent mitigation *M*_*t*_ due to non-pharmaceutical interventions (NPIs), as given in Aguas et al.:

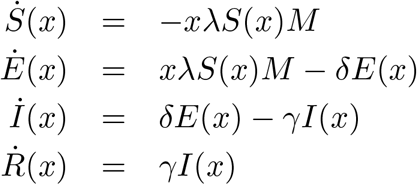

Then,

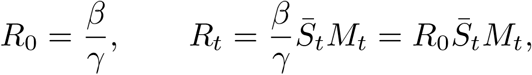

where *R*_0_ is the basic reproduction number, *R*_*t*_ = *R*_eff_(*t*) is the effective reproduction number at time *t*, and 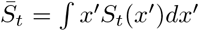 is the mean susceptibility at time *t*. More generally, we can consider time-dependent effective reproduction numbers of the form

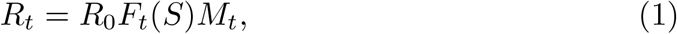

where *F*_*t*_(*S*) is some functional of the variable susceptibility *S*_*t*_(*x*) as a function of *x* (notice that the functional itself does not change over time, but that the resulting function has a time dependence since *S*_*t*_(*x*) does). The model of Aguas et al. corresponds to the case when 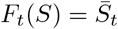. From this equation it is already clear that given a time series *R*_*t*_, we can for every *t* only infer the combined impact of heterogeneity and mitigation (the product *F*_*t*_(*S*)*M*_*t*_), but not each separately.

Now consider 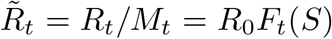, the effective reproduction number if there wasno intervention at all (i.e., if *M*_*t*_ *≡* 1). Herd immunity is reached at 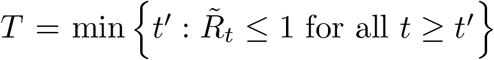 (i.e., the first time after which 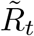 remains at or below 1), and the corresponding herd immunity threshold is

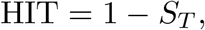

where *S*_*t*_ = ∫*S*_*t*_(*x ′*)*dx*′.

#### Epidemic dynamics with Gamma-distributed variable susceptibility

Following a similar derivation as that in Montalban et al. [11], it can be shown that

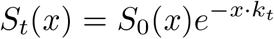

where 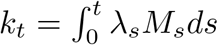 and *λ*_*t*_ is the force of infection at time *t*. Assuming *S*_0_(*x*) is a Gamma(*a, a*) density (where *a* is related to the coefficient of variation by 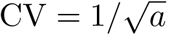, we have that

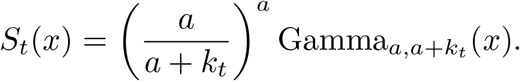

Taking integrals, it can be shown that

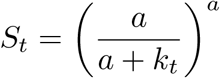

and

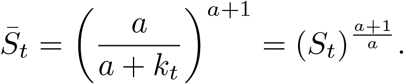

Using the previous formula, we have that 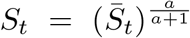 Since 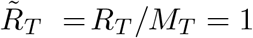, we have that 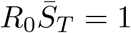
and

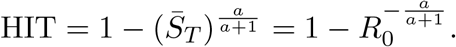

#### Deriving mitigation curves from herd immunity thresholds

We now show how it is possible to derive a mitigation curve *M*_*t*_ that is consistent with the dynamics under Gamma-distributed variable susceptibility for a given herd immunity threshold.

To begin, let us reparameterize time in terms of the proportion of susceptibles *S*_*t*_ to make our calculations simpler. First, Note that *S*_*t*_ is strictly monotone decreasing so long as *λ*_*t*_*M*_*t*_ *>* 0 for all *t* (i.e., both the force of infection and mitigation curve are strictly positive), and that *S*_0_ = 1 and *S*_*t*_ *→* 0 as *t → ∞*. Let *σ*_*t*_ = 1 *− S*_*t*_. Then, we can reparameterize time by *t → σ* (i.e., [0, *∞*) [0, 1]). That is, Equation (1) becomes

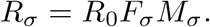

Assume that *F*_*t*_ and *R*_*t*_ are given. Then *M*_*σ*_ can be calculated as

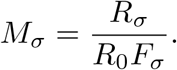

Let us write script letters for log of the values:

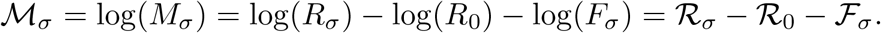

Since *F*_*t*_(*S*) = (*S*_*t*_)^(*a*+1)*/a*^, we have that *F*_*σ*_ = (1 *− σ*)^(*a*+1)*/a*^. Therefore, 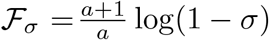 and

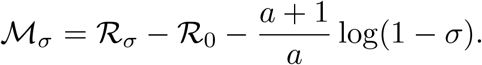

As before, herd immunity is reached at the value *S*_*∗*_ of *S* at which no growth occurs in the absence of intervention, i.e., *R*_*t*_*/M*_*t*_ *≡ R*_*∗*_*/M*_*∗*_ = 1. That is,

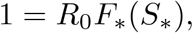

and therefore 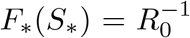. Let *σ*_*∗*_ = 1 *− S*_*∗*_ = HIT. Since *F* (*S*) = *S*^(*a*+1)*/a*^, this implies

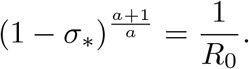

Taking logarithms, we find that

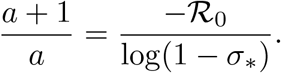

Finally, substituting this into the expression above above we get

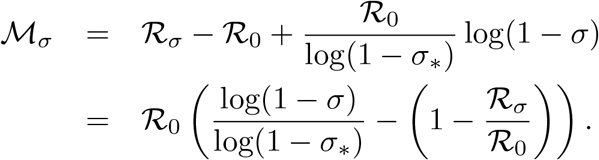

Inverting the mapping *t* ↦ *σ*, we can again write this as a function of time *t*:

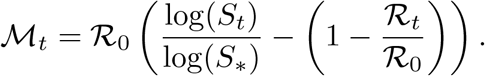

Exponentiating both sides, we finally arrive at the expression

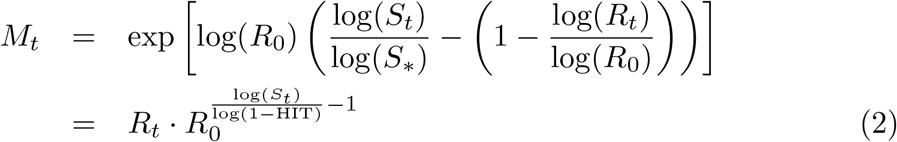

This gives us a formula for calculating a mitigation curve *M*_*t*_ that is consistent with the underlying variable susceptibility model of Aguas et al., for any chosen value of a herd immunity threshold.

#### Computing mitigation curves as a function of herd immunity threshold

We can see from the previous section that the model is attempting to match the dynamics seen in the data through a combination of mitigation and herd immunity as impacted through population heterogeneity. To further show why strong mitigation assumptions are needed to estimate the herd immunity thresholds, we show how one can craft mitigation curves to match dynamics for a wide range of herd immunity thresholds for each of the four countries investigated (Belgium, England, Portugal, and Spain).

Using Equation (2), we can estimate daily mitigation impacts for a given herd immunity threshold (HIT) given the necessary data. To do so, one only needs estimates for the basic reproduction number (*R*_0_), daily effective reproduction number (*R*_*t*_), and the proportion of the population still susceptible to the disease (*S*_*t*_). For each of the four countries we first estimated *R*_*t*_ using the available case data and a common method for measuring the instantaneous reproduction number assuming a mean serial interval of 5.8 days and standard deviation of 4.48 days [4, 8]. We took the largest value of *R*_*t*_ as our rough estimate for the *R*_0_. Finally, we used the case data and the estimated country-specific reporting rates estimated in Aguas et al. to estimate the true incidence of disease each day, which we converted to *S*_*t*_ using *S*_*t*_ = *S*_*t−*1_ *− C*_*t*_*/ρ*. Here, *C*_*t*_ is the number of the reported cases in that country at time *t*, and *ρ* is the country-specific reporting rate from Aguas et al: Belgium (0.06), Portugal (0.09), Spain (0.059), England (0.024).

In Figure S5 we show the calculated mitigation curves for each country across a range of potential HITs from 20% to 70%.

---

**Figure S5:**
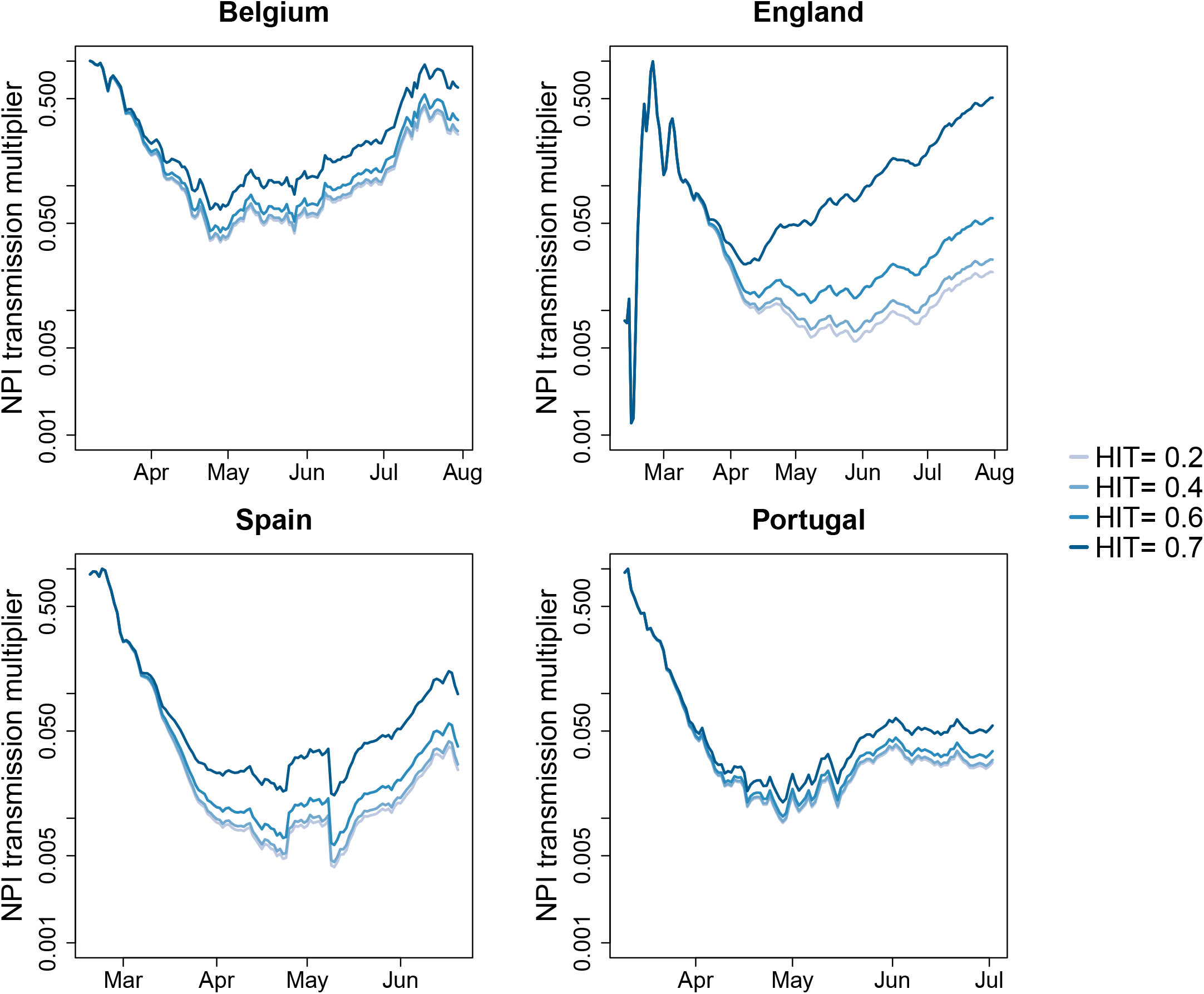
Using cases data and *R*_*t*_ estimation we calculate *M*_*t*_, the NPI transmission multiplier timelines that would be inferred to have a predetermined HIT.

## References

1 Fontanet A, Cauchemez S. COVID-19 herd immunity: where are we? Nat Rev Immunol 2020; 20: 583–4.

2 Aguas R, Corder RM, King JG, Goncalves G, Ferreira MU, M. Gomes MG. Herd immunity thresholds for SARS-CoV-2 estimated from unfolding epidemics. Epidemiology. 2020; published online July 24. DOI:10.1101/2020.07.23.20160762.

3 Keaten J. WHO: 10% of world’s people may have been infected with virus. Associated Press. 2020; published online Oct 5. https://apnews.com/article/virus-outbreak-archive-united-nations-54a3a5869c9ae4ee623497691e796083 (Accessed Nov 10, 2020).

4 Great Barrington Declaration. https://gbdeclaration.org/ (Accessed Nov 3, 2020).

5 Alwan NA, Burgess RA, Ashworth S, et al. Scientific consensus on the COVID-19 pandemic: we need to act now. Lancet 2020; 396: e71–2.

6 Colombo M, Mellor J, Colhoun HM, M. Gomes MG, McKeigue PM. Trajectory of COVID-19 epidemic in Europe. Infectious Diseases (except HIV/AIDS). 2020; published online Sept 28. DOI:10.1101/2020.09.26.20202267.

7 Buss LF, Prete CA Jr, Abrahim CMM, et al. COVID-19 herd immunity in the Brazilian Amazon. Infectious Diseases (except HIV/AIDS). 2020; published online Sept 21. DOI:10.1101/2020.09.16.20194787.

8 Hagan LM. Mass Testing for SARS-CoV-2 in 16 Prisons and Jails — Six Jurisdictions, United States, April–May 2020. MMWR Morb Mortal Wkly Rep 2020; 69. DOI:10.15585/mmwr.mm6933a3.

9 McMichael TM, Currie DW, Clark S, et al. Epidemiology of Covid-19 in a Long-Term Care Facility in King County, Washington. N Engl J Med 2020; 382: 2005–11.

10 Gomes MGM, Corder RM, King JG, et al. Individual variation in susceptibility or exposure to SARS-CoV-2 lowers the herd immunity threshold. medRxiv 2020; published online May 2. DOI:10.1101/2020.04.27.20081893.

11 Hale T, Webster S, Petherick A, Phillips T, Kira B. Oxford covid-19 government response tracker. Blavatnik School of Government 2020; 25.

## References

[1] Ricardo Aguas et al. “Herd immunity thresholds for SARS-CoV-2 estimated from unfolding epidemics”. In: medRxiv (2020). doi: 10.1101/2020.07.23.20160762. url: https://www.medrxiv.org/content/early/2020/08/31/2020.07.23.20160762.

[2] Tom Britton, Frank Ball, and Pieter Trapman. “A mathematical model reveals the influence of population heterogeneity on herd immunity to SARS-CoV-2”. en. In: Science 369.6505 (Aug. 2020), pp. 846–849.

[3] Lewis F Buss et al. “COVID-19 herd immunity in the Brazilian Amazon”. Sept. 2020.

[4] Anne Cori et al. “A New Framework and Software to Estimate Time-Varying Reproduction Numbers During Epidemics”. In: American Journal of Epidemiology 178.9 (Sept. 2013), pp. 1505–1512. issn: 0002-9262. doi: 10.1093/aje/kwt133. eprint: https://academic.oup.com/aje/article-pdf/178/9/1505/17341195/kwt133.pdf. url: https://doi.org/10.1093/aje/kwt133.

[5] Arnaud Fontanet and Simon Cauchemez. “COVID-19 herd immunity: where are we?” en. In: Nat. Rev. Immunol. 20.10 (Oct. 2020), pp. 583–584.

[6] M Gabriela M Gomes et al. “Individual variation in susceptibility or exposure to SARS-CoV-2 lowers the herd immunity threshold”. In: medRxiv (2020). doi: 10.1101/2020.04.27.20081893. url: https://www.medrxiv.org/content/early/2020/05/21/2020.04.27.20081893.

[7] Thomas Hale et al. “Oxford covid-19 government response tracker”. In: Blavatnik School of Government 25 (2020).

[8] Xi He et al. “Temporal dynamics in viral shedding and transmissibility of COVID-19”. en. In: Nat. Med. (Apr. 2020).

[9] Kin On Kwok et al. Herd immunity – estimating the level required to halt the COVID-19 epidemics in affected countries. 2020.

[10] Gideon Meyerowitz-Katz and Lea Merone. “A systematic review and meta-analysis of published research data on COVID-19 infection-fatality rates”. en. In: Int. J. Infect. Dis. (Sept. 2020).

[11] Antonio Montalbán, Rodrigo M. Corder, and M. Gabriela M. Gomes. Herd immunity under individual variation and reinfection. 2020. 2008.00098 [q-bio.PE].

[12] P Nouvellet et al. “Report 26: Reduction in mobility and COVID-19 transmission”. In: (). url: https://spiral.imperial.ac.uk/handle/10044/1/79643.

